# Phenotypic associations with the *HMOX1* GT(n) promoter repeat in European populations

**DOI:** 10.1101/2022.04.28.22274424

**Authors:** Fergus Hamilton, Ruth Mitchell, Peter Ghazal, Nic Timpson

**Author notes:** <u>Corresponding author:</u> Dr Fergus Hamilton, MRC Integrative Epidemiology Unit, Oakfield House, Oakfield Grove, Bristol BS8 2BN.

## Abstract

HO-1 is a key enzyme in the management of heme in humans. A GT(n) repeat length in the gene *HMOX1*, has previously been widely associated with a variety of phenotypes, including susceptibility and outcomes in diabetes, cancer, infections, and neonatal jaundice. However, studies are generally small and results inconsistent. In this study, we imputed the GT(n) repeat length in two European cohorts (UK Biobank, n = 463,005; and Avon Longitudinal Study of Parents and Children (ALSPAC n = 937), with the reliability of imputation tested in other cohorts (1000 Genomes, HGDP, and UK-PGP). Subsequently, we measured the relationship between repeat length and previously identified associations (diabetes, COPD, pneumonia and infection related mortality in UK Biobank; neonatal jaundice in ALSPAC) and performed a phenome-wide association study (PheWAS) in UK Biobank. Despite high quality imputation (correlation between true repeat length and imputed repeat length >0.9 in test cohorts), no clinical associations were identified in either the PheWAS or specific association studies. These findings were robust to definitions of repeat length and sensitivity analyses. Despite multiple smaller studies identifying associations across a variety of clinical settings; we could not replicate or identify any relevant phenotypic associations with the *HMOX1* GT(n) repeat.

## Introduction

The gene *HMOX1*, which encodes for the protein HO-1, is a critical component of life. *HMOX1* is a key enzyme in the heme breakdown pathway, and catalyses the breakdown of heme, an iron containing porphyrin, into bilverdin, Fe2+, and carbon monoxide. As heme is such a critical component of multiple biological systems, with a particular key role in maintaining cellular stress alterations in this gene’s function are suspected to have significant clinical manifestations.^1^

Supporting this, *HMOX1* missense and nonsense mutations in humans are remarkably rare, with less than ten cases reported across the literature, all of which had dramatic phenotypic associations with an increased inflammatory state, liver dysfunction, and marked iron dysregulation.^2^ A large volume of laboratory and animal work has identified the importance of this gene, with animal studies suggesting that up and/or down regulation of this gene has significant impacts on conditions as diverse as malaria,^3,4^ sepsis,^5^ and diabetes.^6^

More than thirty years ago, a GT(n) repeat in the promoter of this gene was identified, that varies from around 15 to 40 copies. Over time, multiple studies have identified clinical manifestations associated with the length of this repeat (last reviewed in 2004, by Exner^1^). Although not all studies have identified robust associations, meta-analyses have identified associations in a variety of conditions such as diabetes,^7^ chronic obstructive pulmonary disease (COPD),^8^ and neonatal jaundice.^9^ In cancer, two meta-analyses suggested a potential role for the *HMOX1* GT(n) repeat in susceptibility in Asian populations. ^10,11^

Further laboratory work has confirmed the functionality of this repeat, with most, but not all studies identifying that this repeat alters the inducible expression of *HMOX1*.^12–18^ Therefore, in this study, we aimed to identify phenotypic associations with the *HMOX1* promoter repeat in two European cohorts: UK Biobank and ALSPAC, by using a recently developed imputation reference panel including this repeat.^19^

In particular we performed three specific analyses: Firstly, we searched for any phenotypic associations across UK Biobank in a Phenome wide association analysis (PheWAS), then focussing on two previously reported associations: infections and neonatal jaundice.

## Methods

### UK Biobank access, genetic data, and quality control

#### UK Biobank

UK Biobank is a population-based health research resource consisting of approximately 500,000 people, aged between 38 years and 73 years, who were recruited between the years 2006 and 2010 from across the UK.^20^ Particularly focused on identifying determinants of human diseases in middle-aged and older individuals, participants provided a range of information (such as demographics, health status, lifestyle measures, cognitive testing, personality self-report, and physical and mental health measures) via questionnaires and interviews; anthropometric measures, BP readings and samples of blood, urine and saliva were also taken (data available at www.ukbiobank.ac.uk). A full description of the study design, participants and quality control (QC) methods have been described in detail previously.^20,21^ UK Biobank received ethical approval from the Research Ethics Committee (REC reference for UK Biobank is 11/NW/0382). This study was performed under application number 52643.

#### Genotyping and imputation

The full data release contains the cohort of successfully genotyped samples (n=488,377). 49,979 individuals were genotyped using the UK BiLEVE array and 438,398 using the UK Biobank axiom array. Pre-imputation QC, phasing and imputation are described elsewhere.^20^ In brief, prior to phasing, multiallelic SNPs or those with MAF ≤1% were removed. Phasing of genotype data was performed using a modified version of the SHAPEIT2 algorithm. Genotype imputation to a reference set combining the UK10K haplotype and HRC reference panels was performed using IMPUTE2 algorithms. The analyses presented here were restricted to autosomal variants using a graded filtering with varying imputation quality for different allele frequency ranges. Therefore, rarer genetic variants are required to have a higher imputation INFO score (Info>0.3 for MAF >3%; Info>0.6 for MAF 1-3%; Info>0.8 for MAF 0.5-1%; Info>0.9 for MAF 0.1-0.5%) with MAF and Info scores having been recalculated on an in-house derived ‘European’ subset.^21^

#### Data quality control

Individuals with sex-mismatch (derived by comparing genetic sex and reported sex) or individuals with sex chromosome aneuploidy were excluded from the analysis (n = 814). Individuals who were outliers in heterozygosity and missing rates (n = 968).

We restricted the sample to individuals of ‘European’ ancestry as defined by an in-house k-means cluster analysis performed using the first 4 principal components provided by UK Biobank in the statistical software environment R. The current analysis includes the largest cluster from this analysis (n = 464,708).

#### ALSPAC

ALSPAC is a longitudinal birth cohort that recruited 14 541 pregnant women resident in Avon, UK, with expected dates of delivery between 1 April 1991 and 3 December 1992. Of these initial pregnancies, there were a total of 14 676 fetuses, resulting in 14 062 live births and 13 988 children who were alive at 1 year of age. The children and their mothers have been followed up through postal questionnaires and at clinics.^22,23^ Please note that the study website contains details of all the data that is available through a fully searchable data dictionary and variable search tool.^24^ Ethical approval for the study was obtained from the ALSPAC Ethics and Law Committee and the Local Research Ethics

#### Committees (ref B3801)

ALSPAC children were genotyped using the Illumina HumanHap550 quad chip genotyping platforms, and genotypes were called with Illumina GenomeStudio. Imputation was performed using the Michigan Imputation Server with phasing via SHAPEIT. SNP-level quality control removed variants with more than 5% missingness, or p value for Hardy–Weinberg equilibrium smaller than 1e−6. Participant-level quality control removed variants with uncertain X chromosome heterozygosity, extreme autosomal heterozygosity or more than 5% overall missingness. Next, multi-dimensional scaling of genome-wide data was performed including reference data from HapMap populations. Samples which clustered outside the CEU population were removed. We limited SNPs to those with an INFO score of >0.8.

#### Other cohorts

For testing the robustness of the imputation, whole genome and SNP array data was downloaded from three further datasets: The Human Genome Diversity project (HGDP),^25^ The UK Personal Genome Project (PGP),^26^ and the 1000 Genomes Project (1KGP).^27^ All of these datasets are freely available without prior ethical approval.

#### Imputation approach

An imputation reference panel for short tandem repeats has previously been developed for use with SNP array data.^19^ This was developed using linked SNP array/WGS data from two cohorts (Simon’s Simplex Cohort and 1000 Genomes). The HipSTR tool was used to call repeat length from WGS data, and then a reference panel was developed for use in downstream imputation.^28^

For this study, genomic data for a 2mb region either side of *HMOX1* were extracted from the above datasets, and lifted over to the GrCh37 reference, if not on it already. Alleles were conformed using conform-gt to ensure they were the same as the reference genome, and then Beagle 5.2 was used with standard settings with a window of 2mb to impute the *HMOX1* STR, using the previously developed reference panel.^19^ Only high quality SNPS (INFO > 0.8) were included.

#### Allelic calling and genotyping

As the repeat length varies in a linear fashion, the main analysis was done using the sum of both allelic lengths as a linear predictor of phenotypes. The length was calculated from the number of full GT repeats. We chose to include the repeat as a linear approach (as opposed to looking at each individuals haplotype) because a) prior associations at this repeat have always been linear, and b) nearly all previously published STR associations have been linear.^1,29^

For sensitivity analyses, various allelic definitions were tested, including an allelic model (where each allele was run separately), and a model using repeat length cut-offs to define alleles. There is significant variation in the literature around what defines an appropriate cut off, with both binary (short vs long) and ternary models (short, medium and long), with a wide variety of cut offs used across prior studies. Given this variation; and with no empiric evidence to support previous definitions, we chose to simply quartile the alleles and include them in the model. ^30^

#### Imputation quality control

We tested the imputation quality in four discrete datasets (UKB, PGP, HGDP, 1KGP) with combined high quality (>20x coverage) WGS data and SNP array data available for the same sample. For the WGS data, the locus was called directly from the BAM files using HipSTR v 0.6.2 with standard settings.^31^ Subsequently, standard filtration to remove low quality calls was performed as per HipSTR standard settings, and only samples with a posterior probability of the correct genotype of > 0.9 were kept. Full details of the imputation quality control and testing are in Supplement S2.

No quality control was performed on the imputed data, as initial experimentation identification identified that the posterior probability of genotype call was not predictive of accuracy (Supplement S2)

The quality of imputation at this locus using the haplotype reference panel has previously been reported to be good at this locus, particularly in European populations.^32^ Three imputation metrics are reported, the concordance (the fraction that exactly match the allele length across both datasets), the Pearson’s correlation (R), calculated on the sum across both alleles, and the fraction concordant to within two repeats, as most previous analysis have suggested a linear relationship between STR length and outcomes.^29^ As a final test of imputation, we imputed the AC promoter repeat in *RFT1* in UK Biobank. This repeat has been previously associated with height in the eMERGE cohort.^29^

#### UK Biobank PheWAS

Using the above genotypes for *HMOX1*, a PheWAS was performed across the UK Biobank participants for a wide variety of traits using an established software (PHESANT).^33^ Traits included: Algorithmically defined health outcomes extracted from EHR data (e.g. diagnoses), anthropometric traits, biological sample traits (e.g. protein levels), health questionnaires, and mortality data. In total, 7,901 traits were included. Full details of the pipeline are available in the original publication, but briefly, for linear traits, linear regression was used, for binary traits, logistic regression was performed, for ordered categorical traits, ordinal logistic regression was used, and for unordered traits, multinomial regression was used. The analysis was run using age, sex, genetic chip, and the first 10 principal components as covariates. A complete case analysis was performed for each trait (i.e. no imputation).

As a sensitivity analysis, the analysis was without any covariates, and using repeat length split into equal groups to explore any non-linear effects. Previous studies have used a variety of cut-offs, with no clear evidence to support any particular allelic definition. Therefore, we simply quartiled the exposure and re-ran the analysis.

#### UK Biobank infection specific analyses

As *HMOX1* is a stress response gene, it is plausible that the impact of any genetic variation is only present in the presence of cellular stress; so in a cohort type analysis, no signal of variation would be identified, despite a signal during cellular stress. In particular, infection has been suggested as a particular cellular stress and severe infection is known to highly upregulate *HMOX1*, while knockout models of *HMOX1* in animals show markedly worse outcomes with infection.

Therefore, we extracted cases of infection from UK Biobank and used Cox regression to estimate hazards for 28-day mortality, with *HMOX1* allele length as a predictor. Definitions of infection were developed by an infectious disease specialist (FH) using ICD-10 coding, and extracted from the UK Biobank linked EHR data.^34^ Codes for each infection are available in the supplement. Cox regression was performed using the survival package in R,^35^ and run both unadjusted and adjusted for age and sex.

As patients often had multiple episodes of infection, for this analysis, only the latest was taken, to avoid immortal time bias.

#### ALSPAC neonatal jaundice

As *HMOX1* promoter variation has been shown to influence rates of neonatal hyperbilirubinemia, data on bilirubin levels around birth and date of sampling were extracted for 937 ALSPAC participants. Definitions of neonatal jaundice vary between governing bodies, with both the time since birth (in hours), and the level of the bilirubin result influencing case definitions. For ALSPAC participants, we had recorded the highest recorded bilirubin result, and the day post birth that this result had occurred.

Therefore, we took two approaches. Firstly, we used the NICE nomogram for bilirubin levels to generate cases of neonatal hyperbilirubinemia, and then performed a case control analysis with the *HMOX1* promoter genotype as a predictor of case status.^36^ As cases of neonatal jaundice were expected to be low in this birth cohort, we then subsequently developed a Z-score for each participant for the given day the sample was taken on, so each participant with a result had a Z-score for the bilirubin result for a given day. Repeat length of the *HMOX1*S TR was then tested as a predictor of Z-score, in a linear fashion.

#### Software

All statistical analysis was performed in R v4.0.4, using the packages: tidyverse, survival, data.table, and gtsummary. Specific genetic analyses are described above with the relevant software.

## Results

### Imputation quality

The quality of the imputation was tested across 4 discrete datasets (1000 Genomes, Human Genome Diversity Project, Personal Genome Project UK, and UK Biobank, Table 1, Figure 1), all of which had both SNP array data and high quality WGS data. In particular, we compared Pearson’s correlation between the total repeat length across both alleles, and the exact concordance (both alleles exactly correct).

**Table 1:**
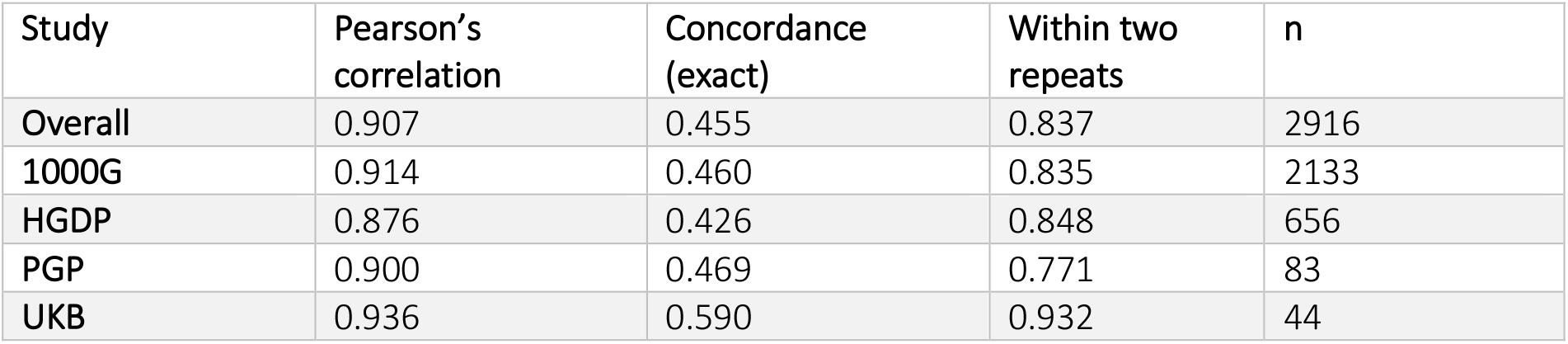
Overall imputation performance across the four test datasets.

**Figure 1:**
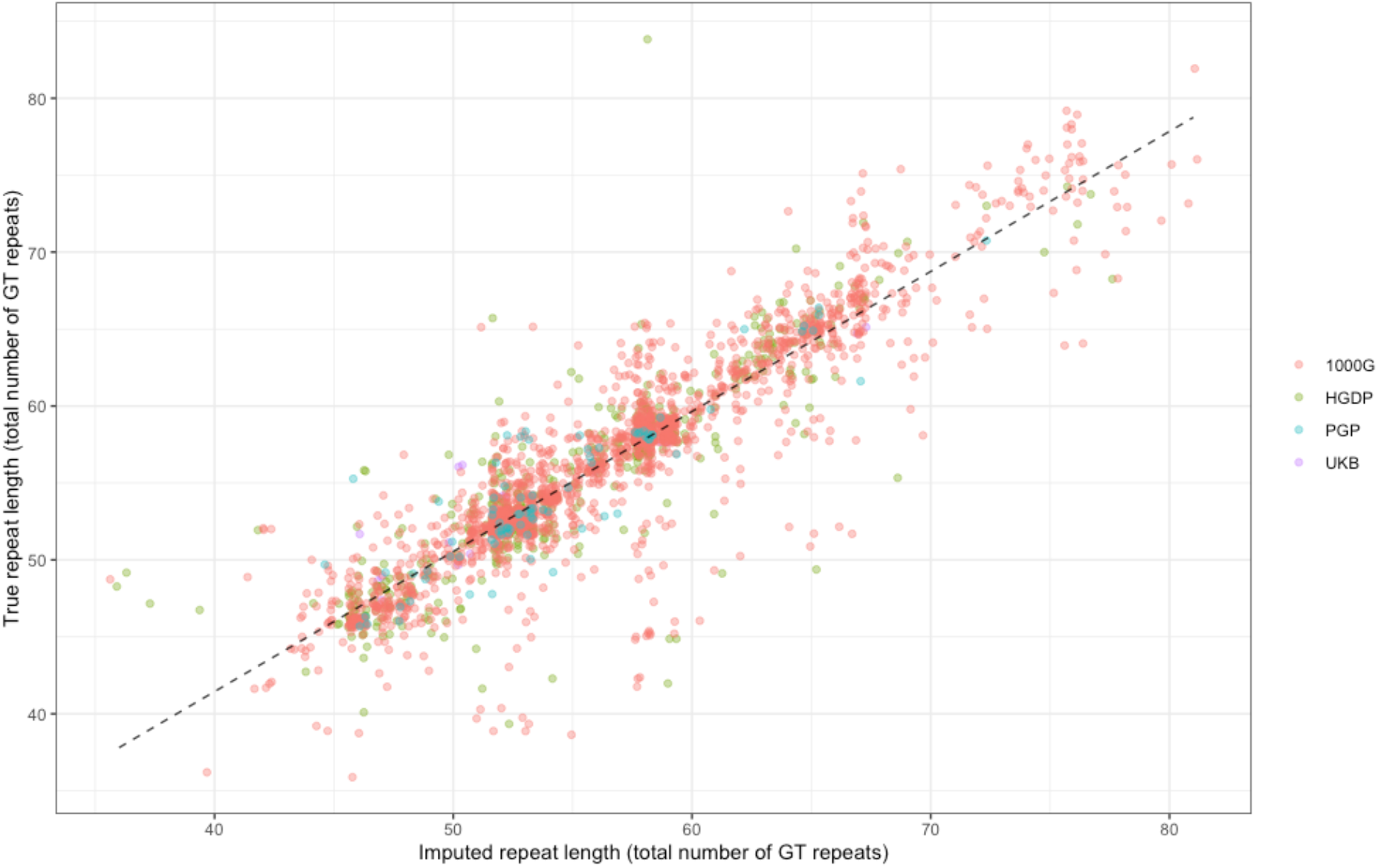
Overall imputation performance at the *HMOX1* STR.

In summary, the correlation between the summed length of both alleles was excellent, with a Pearson’s R of 0.91 in the whole cohort. However, concordance -where both alleles were correctly called to the exact repeat, was much weaker with a summary concordance of 0.45. Importantly, however, >80% of all called lengths were accurate to within two repeats, and >90% across UK Biobank, our main cohort. The median absolute difference in repeat lengths was 1, IQR 0-2, and there was no evidence of systemic positive or negative bias in imputing (mean difference between imputed and true repeat length -0.0013). Figure S1 shows the median absolute difference in total repeat length across the four cohorts.

As there might be evidence that imputation quality differed between populations, given the different reference haplotypes across populations, we calculated the imputation quality for populations, where this was reported in HGDP and 1000G (Supplement S2). As both PGP and UKB are entirely based in the U.K, this analysis was not undertaken for these two cohorts. In summary, there was some evidence that imputation quality differed between populations, with slightly worse performance in the HGDP superpopulations Africa (n = 50, r = 0.73), East Asia (n = 158, r = 0.84) and Oceania (n = 21). Importantly for this study, there was high correlation between the true repeat length and the imputed repeat length in British ancestry populations indicating that the imputation was of high quality, with correlations above > 0.9 from UKB, PGP, and the GBR populations in 1000G and HGDP.

As a final test of imputation in UK Biobank, we imputed an AC repeat in *RFT1* as a positive control, as it has a known association with height in prior analyses.^29^ In Fosting et al; this repeat was imputed in the eMERGE cohort; where a robust positive association with height was identified (p = 0.00328; beta = 0.010; n = 6,393, for each AC repeat copy).

We replicated this analysis in UK Biobank, with 441,832 participants having both genotype and standing height available for analysis. Reassuringly, we found almost the exact same effect size (beta = 0.011, p < 1 × 10^−16^, Figure 2), showing a small increase in height with increasing repeat length.

**Figure 2:**
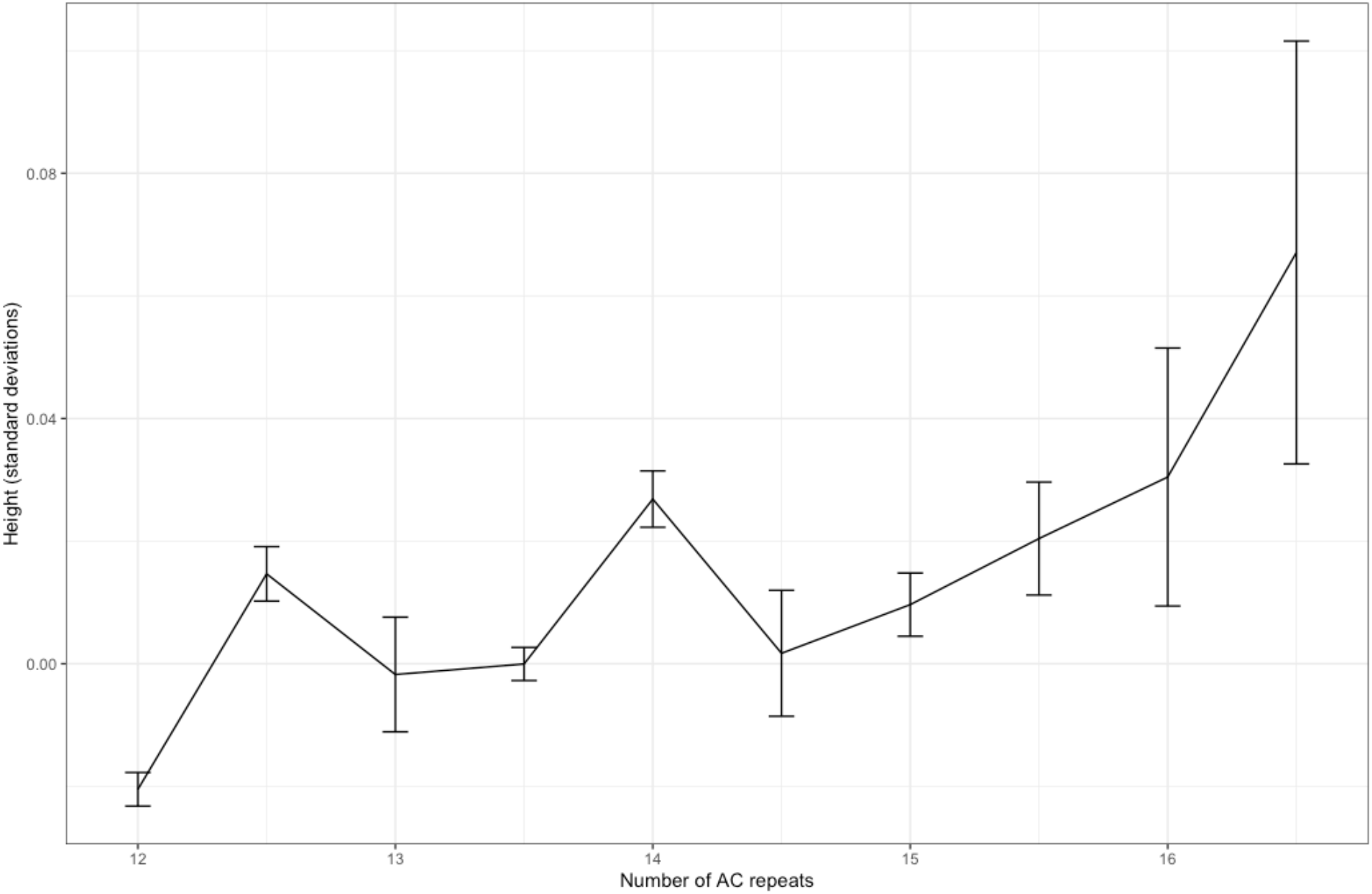
Change in height with increasing AC repeats at RFT1 (showing genotypes where n>500, mean, with bar representing standard error of the mean)

In summary, imputation at the *HMOX1* locus in UK Biobank was reliable (Pearson’s R > 0.9 in British populations), and we were able to replicate other STR-phenotype associations from other cohorts.

### UK Biobank imputation metrics

After quality control and filtering (described above, and here^21^), 463,005 individuals were included in the imputation pipeline. As we performed no filtering post imputation (see Supplement 2 for reasoning), we called *HMOX1* repeat length on 463,005 samples. Figure 3 describe the logged distribution of the allele length for a) individual alleles, and b) summed repeat length across both alleles. We performed a Chi-squared test to compare proportions of homozygotes to heterozygotes at each allele length, which showed that this did not exceed the Hardy Weinberg equilibrium (p = 0.265).

**Figure 3:**
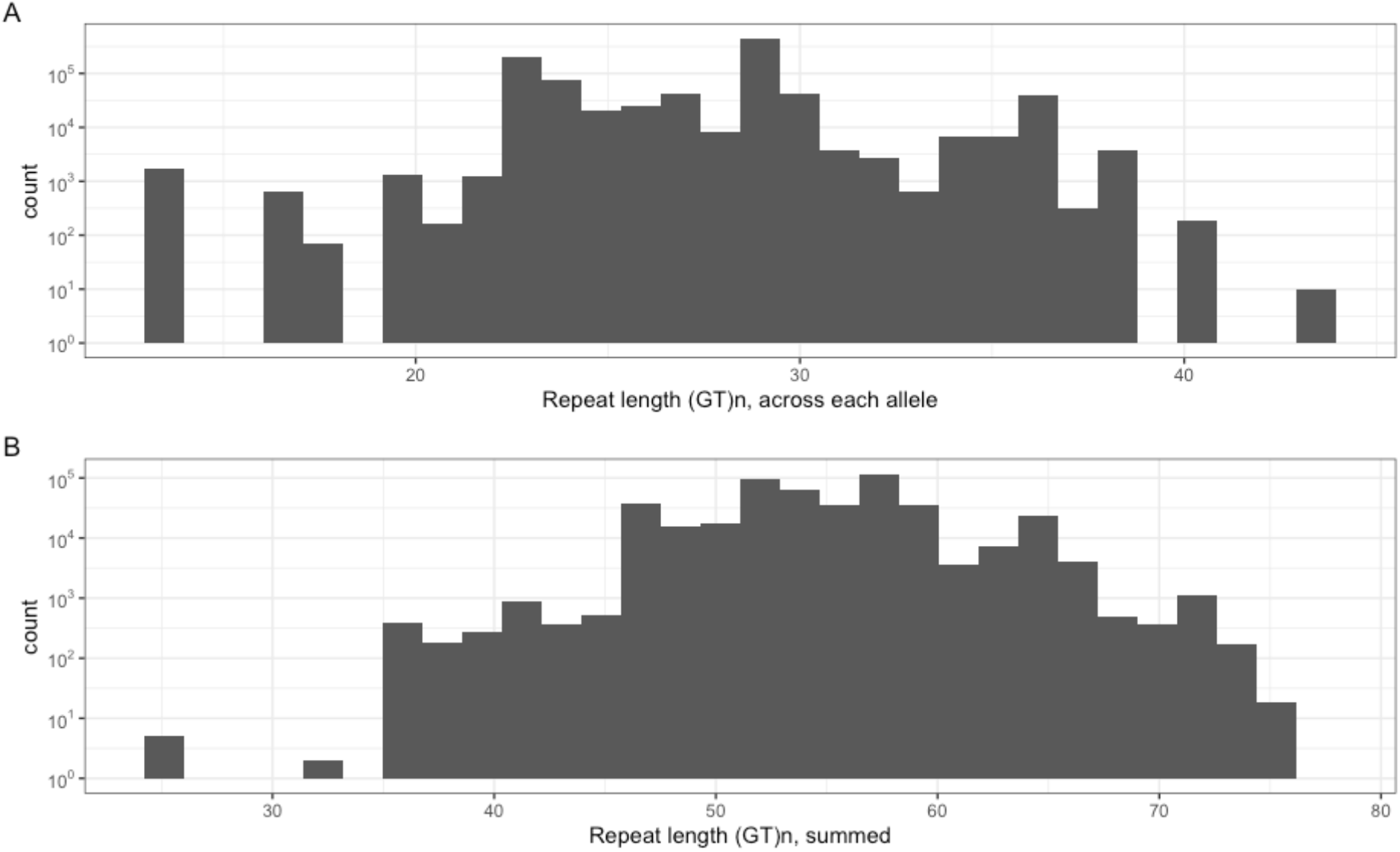
Repeat lengths in UK Biobank from a) individual alleles, and b) summed across both alleles for each individual (log y scale)

As has been shown previously, the *HMOX1* polymorphism has a trimodal distribution, with major peaks at 25, 32, and 39,^19,26^ although in this analysis, the longer repeat length is much rarer, with 32 being by far the most common repeat length.

### PheWAS analysis

For the main PheWAS analysis, we tested 7,901 variables, using a previously described software (PHESANT), and taking *HMOX1* repeat length as a linear variable.^33^ The QQ plot is shown in Figure 4; which shows limited deviation only at the extreme end. No associations met either a Bonferroni adjusted or FDR corrected p-value.

**Figure 4:**
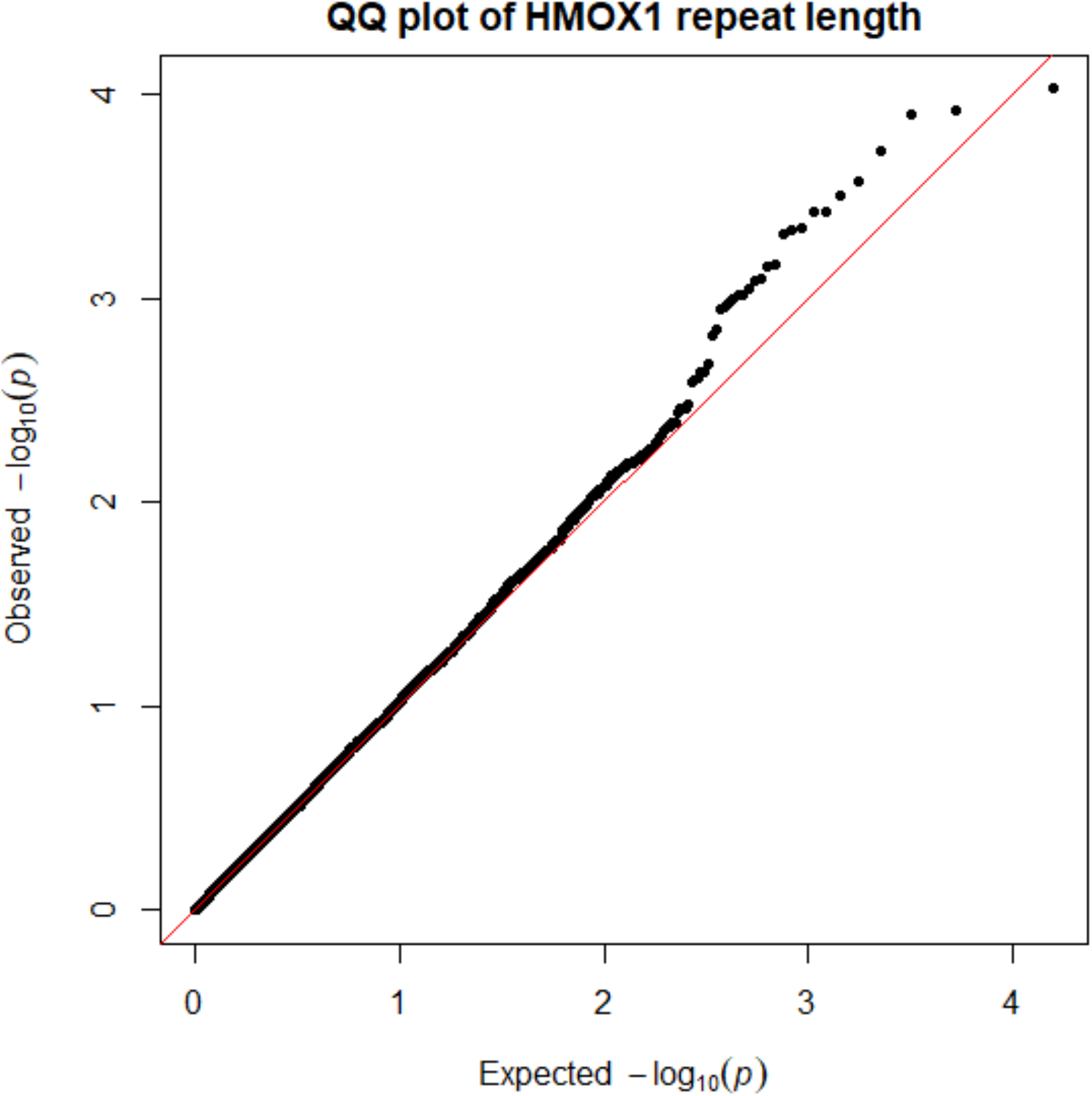
QQ plot of the *HMOX1* PheWAS.

In our sensitivity analyses, we performed the PheWAS without adjustment for principal components or UK Biobank assessment centre and quartiled the exposure, to ensure that our definition of the exposure (summed repeat length) did not alter the results (Supplement S3). For the latter two models, we had similar results to the main model, while in the former model (without adjustment), we identified associations with some sociodemographic variables (e.g. place of birth, month of attending assessment centre), but did not identify any clinically relevant associations.

### Specific associations

As pneumonia, COPD, and diabetes have all previously been identified as having an association with the *HMOX1* promoter repeat, we extracted these clinical variables from the PheWAS analysis. None showed evidence of an association with *HMOX1* repeat length (Table 3).

**Table 3:**
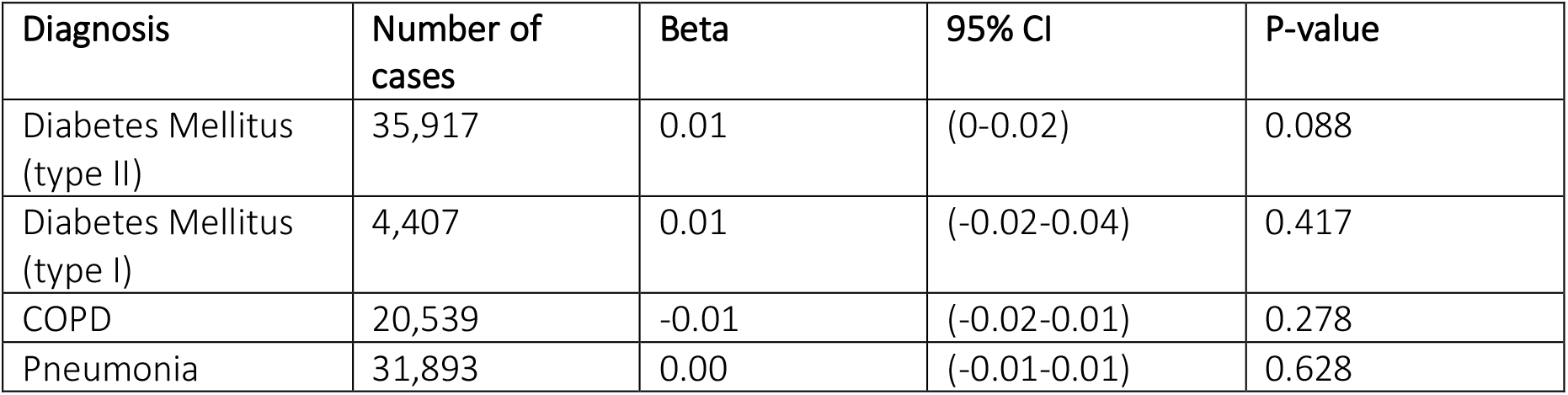
Specific previously identified associations.

| Diagnosis | Number of cases | Beta | 95% CI | P-value |
| --- | --- | --- | --- | --- |
| Diabetes Mellitus (type II) | 35,917 | 0.01 | (0-0.02) | 0.088 |
| Diabetes Mellitus (type I) | 4,407 | 0.01 | (-0.02-0.04) | 0.417 |
| COPD | 20,539 | -0.01 | (-0.02-0.01) | 0.278 |
| Pneumonia | 31,893 | 0.00 | (-0.01-0.01) | 0.628 |

### Mortality within infection analyses

As the impact of *HMOX1* repeat polymorphisms might only be present in specific environmental associations such as in severe cellular stress, and there is some evidence that *HMOX1* polymorphisms alter survival from severe infection,^14,30^ we undertook a survival analysis for 11 coded infections within UK Biobank (Table 4).

**Table 4:** Included infections in UK Biobank, with associated mortality.

| Diagnosis | Number of cases | 28 day mortality (%) |
| --- | --- | --- |
| LRTI | 61864 | 1.66 |
| URTI | 51143 | 0.07 |
| SSTI | 49369 | 0.70 |
| Pneumonia | 35299 | 13.63 |
| Gastroenteritis | 32389 | 2.07 |
| UTI | 17488 | 0.14 |
| Sepsis | 14841 | 16.78 |
| Appendicitis | 10265 | 0.13 |
| Osteomyelitis | 7793 | 1.03 |
| Cholecystitis | 5974 | 1.00 |
| Endocarditis | 1411 | 14.17 |

Infections with a low 28-day mortality were excluded from the analysis and only three infections were taken forward for formal modelling (endocarditis, pneumonia, and sepsis). In both unadjusted and adjusted (for age and sex) models, there was no association between the *HMOX1* repeat length and hazard of 28-day death (Table 5).

**Table 5:** HR for 28-day mortality with increasing repeat length in each condition.

| Diagnosis | Hazard Ratio | 95% CI | P value |
| --- | --- | --- | --- |
| Endocarditis | 1.004 | 0.977-1.03 | 0.76 |
| Endocarditis (adjusted) | 1.004 | 0.977-1.03 | 0.75 |
| Pneumonia | 1.003 | 0.997-1.01 | 0.29 |
| Pneumonia (adjusted) | 1.003 | 0.997-1.01 | 0.36 |
| Sepsis | 1.001 | 0.993-1.01 | 0.87 |
| Sepsis (adjusted) | 1.001 | 0.993-1.01 | 0.83 |

### Neonatal jaundice

Previous systematic reviews have identified an association between the *HMOX1* promoter and neonatal jaundice.^9^ In ALSPAC, a proportion of children had (clinically driven) bilirubin testing during their post-natal care. In total, 937 children had a bilirubin result within 14 days of birth and were successfully genotyped for the *HMOX1* promoter repeat, using the same pipeline as above. Figure 5 shows the sample distribution, with most samples taken in the first 72hrs of life.

**Figure 5:**
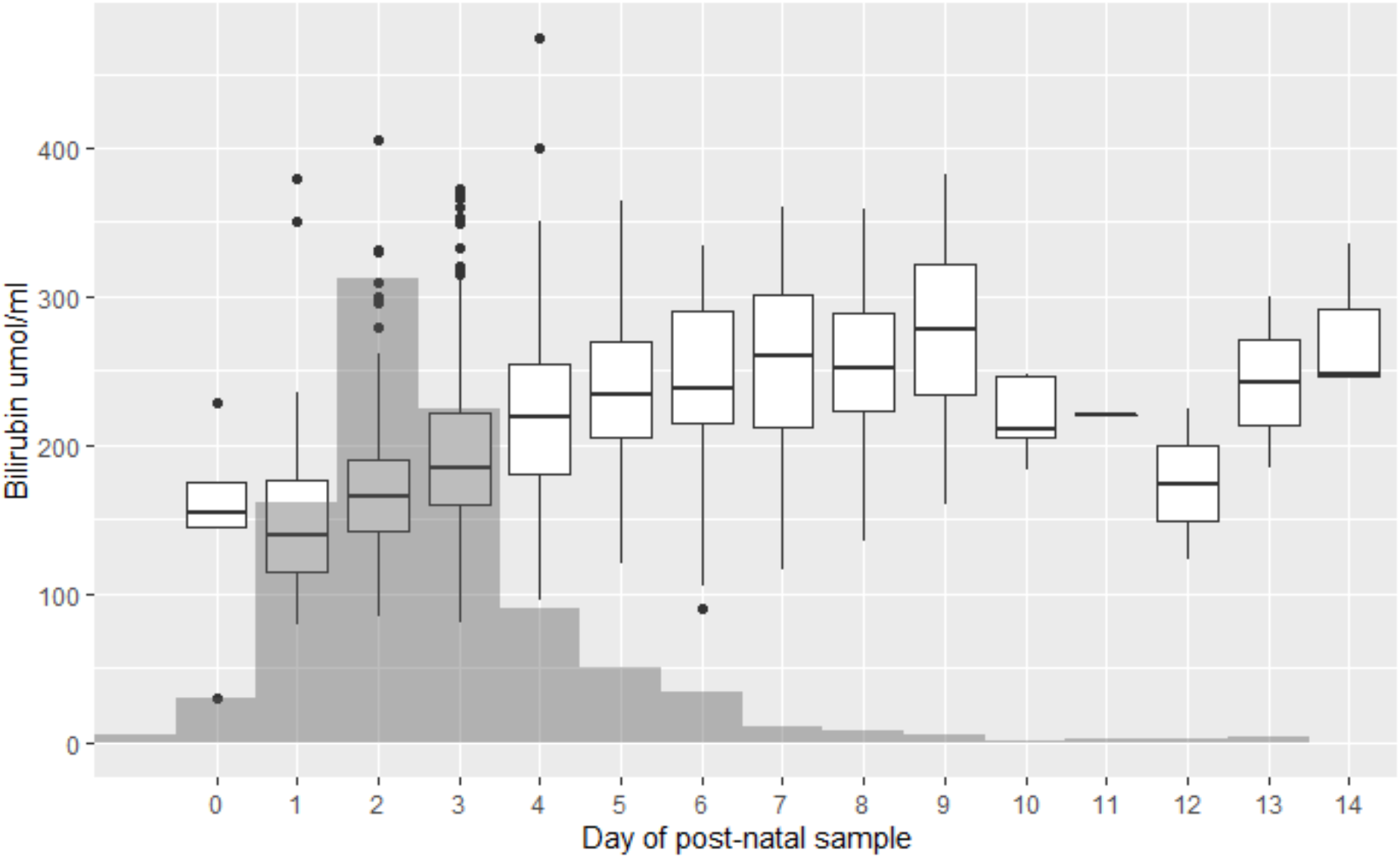
Number of bilirubin samples taken in the ALSPAC cohort, related to post-natal age.

Using the NICE definition of jaundice requiring phototherapy, we calculated the number of cases of neonatal jaundice across our sample. ALSPAC only reports the day of maximal bilirubin, so each child had a single result attached to a single day. ^36^

In total, 47 cases of hyperbilirubinemia were identified, with the vast majority on day 2 (7 cases), 3 (19 cases) or 4 (17 cases) of life. In a logistic regression model, there was no association between total repeat length and neonatal jaundice (OR 1.01; 95% CI 0.88-1.14), and no difference in average repeat length between cases and controls (Mean repeats 27.3 for each group).

Given the low number of cases, we then performed a further analysis, generating a Z-score for each individual result within each day (e.g. each day was treated individually, given the known association between postnatal age and bilirubin level). We then used this Z-score in a linear regression (Figure 6). Again, we identified no association between Z-score and *HMOX1* repeat length (beta < 0.01, p = 0.97).

**Figure 6:**
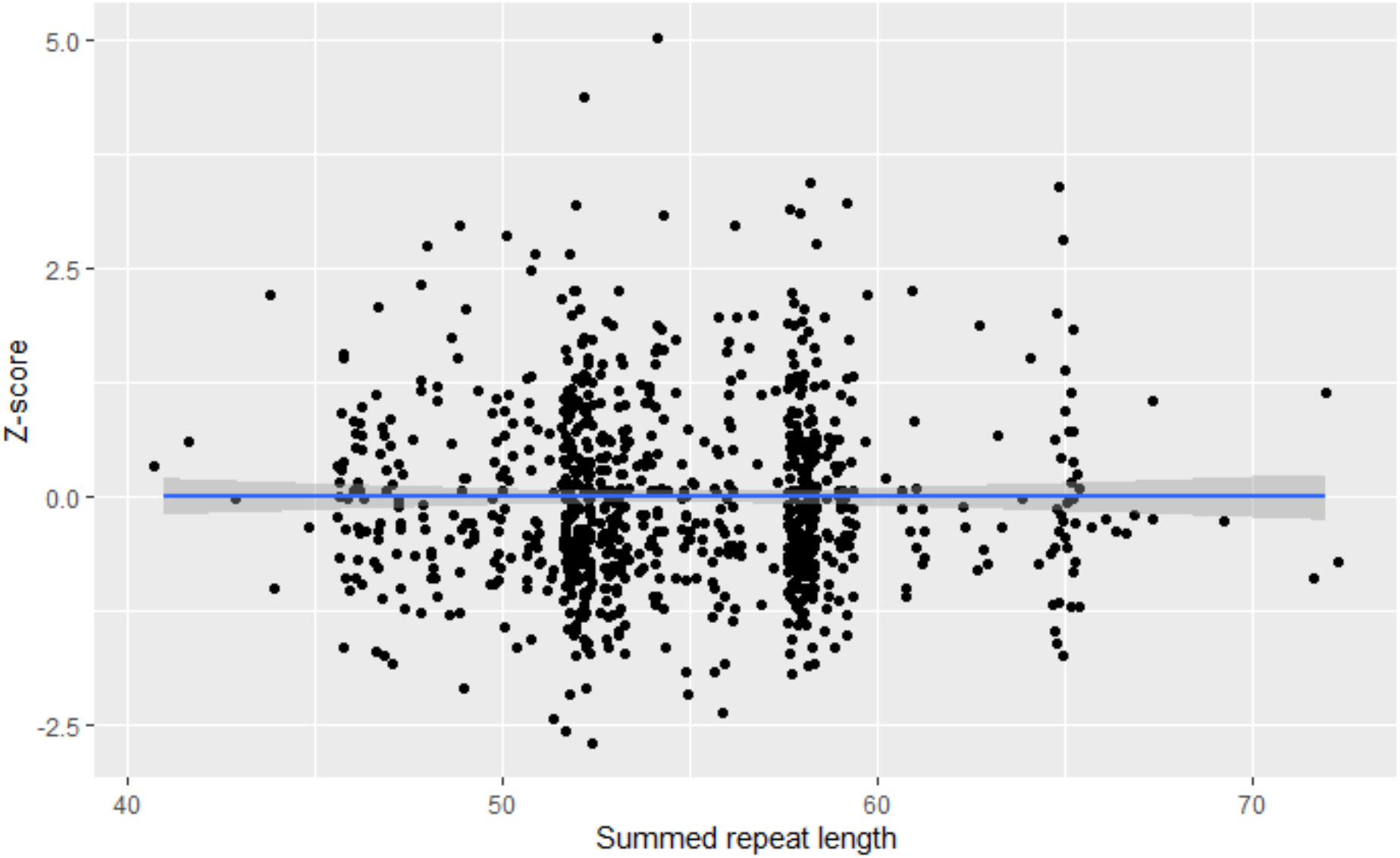
Association between bilirubin Z-score and *HMOX1* repeat length.

In summary, we could not identify any association between the repeat length polymorphism and either clinical jaundice, or an increase in bilirubin levels in neonates, contrary to previous reports.

## Discussion

In this study, we imputed the *HMOX1* repeat polymorphism in two well described cohorts, UK Biobank, and ALSPAC. Imputation accuracy was assessed using external cohorts with high quality WGS data and was generally found to be high (Pearson’s R ∼ 0.9 for imputed repeat length), although concordance with the exact length was lower (around 45%). Using these imputed genotypes, we performed a PheWAS (in UK Biobank), and tested four specific associations from the literature (Pneumonia, COPD, Diabetes, and neonatal jaundice). Finally, we looked to identify if *HMOX1* repeat length was associated with survival from three important infections (Pneumonia, sepsis and endocarditis). In three of these previous conditions (COPD, diabetes, and neonatal jaundice), a meta-analysis of smaller studies has found an association, but we found no associations.

### Strengths

The major strengths of this study are the size of the cohort (for UK Biobank especially), with well characterised clinical metadata. We performed extensive external testing of the imputation approach in four differing cohorts, and found that the imputation accuracy was reliable, particularly in European populations, lending weight to our results. Also, we tested other identified associations with a repeat promoter in RTF1, and were able to replicate others results, suggesting our imputation approach is robust.

### Weaknesses

There are three main weaknesses of this study. The first is that genotypes were imputed, not directly called. However, this is true for many SNPs in most genetic studies, and we have confirmed the reliability of this imputation in four separate datasets; while other published data supports the reliability of the imputation.^36^ Secondly, although the imputed repeat length was highly correlated with true repeat length, it was much less reliable at calling the exact allele length (∼45% correct). This is partly due to a large number of potential alleles (64 potential repeat lengths), with a smaller number of common alleles, making imputation technically hard.

Although this technical limitation should be recognised, it is important to note that all prior associations with this repeat have been with repeat length, with no data that we are aware of suggesting the effect is related to a particular allele rather than the total length of the repeat.^1,7– 10,14,16,30,37^. Secondly, a genome-wide analysis of 2,060 expression short tandem repeats found linear associations to be the most common association between STR’s and gene expression, a finding also identified in other studies of human STRS.^29,38–40^ We therefore suggest that the lack of association with repeat length and multiple phenotypic outcomes in this analysis is valid.

Finally, in common with all cohort analyses, we are limited by the sample frame of the population, which does not represent the wider UK population, and are limited in the outcomes that are recorded, which are largely algorithmically defined from linked electronic health data, or via self-report. However, the quality of linked electronic data in UK Biobank is excellent, and it has been used widely in PheWAS analyses.^41^

### Comparison with previous literature

It is worth exploring the discrepancy between our results and others, particularly in neonatal jaundice and in diabetes, where the strongest prior evidence for an association has been identified. There are multiple possible explanations which are not all mutually exclusive. The most simple explanation is that the null result is correct. The previous studies are all much smaller, with most studies comprising under 1000 patients, with potential biases due to selection of controls, definition of alleles, or the genotyping process, and the evidence for any effect is only present in meta-analyses. It is well established that genetic effects are generally small, and this may simply reflect the common procedure of larger studies identifying null results while smaller, earlier studies, suggest an association.

However, other possible explanations are plausible. Nearly all previous studies have classified alleles into short, medium or long, based on a variety of classification systems. In a previous review of *HMOX1* promoter polymorphisms and infection, we showed that this classification was inconsistent, and creates a significant risk of bias.^30^ For example, in the previous meta-analysis on type 2 diabetes, one study was excluded as allelic definition was too different from the other studies, while they called short alleles as under 25-27 repeats, and long alleles as 25 to 27 repeats.^7^ However, given that i) the trimodal distribution peaks at 27 repeats, ii) definitions of repeat length vary and iii) there are generally small errors and variability in genotyping by fragment length polymorphism, these definitions are highly questionable and are at significant risk of biasing studies. It may be that by selecting allelic definitions, artificial associations were identified.

Secondly, our study was performed entirely in a European population, whereas many of the previous positive studies have been performed in other populations, particularly in East Asia and Africa. As the *HMOX1* repeat promoter association may not be causal, but simply co-incident with local SNP haplotype, it may be that there is an association between certain SNPs in the HMOX1 promoter and outcomes, that are simply not present at a high enough frequency in a European population. Recent studies have suggested that variable number tandem repeats have differing effects on differing haplotypic backgrounds, although this has not been shown for *HMOX1*.^42^

Thirdly, although we tested our imputation approach extensively, and are confident of the accuracy to within 2 repeats, it may be that the imputation is biased with respect to important outcomes. This may be particularly relevant if for example, a set of SNPs that are causal for the outcomes and are usually coincident with an increased repeat length are systematically imputed incorrectly. Although this association is possible, it would be extremely unlikely; as given the number of cases in UK Biobank, we would have ample power to detect even attenuated exposures, if the “true” effect size was similar to that in previous studies.

Finally, although we are able to rule out associations between *HMOX1* repeat length and incident conditions such as diabetes, we cannot easily explore the association between an inducible stress response gene and relevant outcomes (e.g. infection outcomes) in a biobank analysis. In previous *in-vitro* analyses the baseline expression of HMOX1 is unchanged with differing repeat lengths. however, inducible expression is highly varied in some (but not all) in-vitro work.^12,14,16,43^ This suggests that the effect of *HMOX1* promoter polymorphisms may only be present under certain phenotypic conditions (e.g. cellular stress secondary to inflammation or infection). Even given the size of UK Biobank, it is difficult to explore these potential interactions.

For example, if *HMOX1* repeat length only has phenotypic implications in the critically unwell patients with infection, we are limited to those patients in UK Biobank who develop critical infection – an extremely heterogenous condition -while the only outcome measure we can reliably record is mortality.

### Summary

In summary, we did not identify any associations between the *HMOX1* repeat length polymorphism and any clinical outcomes in two well characterised European cohorts. Reconciling this work with previous work is difficult, suggesting either a no association or a more complex gene vs environement interaction for the *HMOX1* repeat.

### Conclusion

The *HMOX1* GT(n) repeat was not associated with any phenotypes in UK Biobank. Previous associations with diabetes, COPD, and pneumonia were not replicated. In a separate analysis, *HMOX1* GT(n) repeat length was also not associated with neonatal jaundice in a longitudinal cohort, failing to replicate previous findings.

## Supporting information

STREGA

## Data Availability

Data is available via the UK Biobank and ALSPAC IDAC.

## Acknowledgments

We are extremely grateful to all the families who took part in the ALSPAC study, the midwives for their help in recruiting them, and the whole ALSPAC team, which includes interviewers, computer and laboratory technicians, clerical workers, research scientists, volunteers, managers, receptionists and nurses. We also thank the participants and staff of UK Biobank.

