## Supplement S1 for "Phenotypic associations with the *HMOX1* GT(n) promoter repeat in European populations"

**Table S1: ICD codes associated with each infection**

| Condition | ICD codes |
| --- | --- |
| Sepsis | A02, A39, A40, A41 |
| Endocarditis | I33, I38, I39 |
| Pneumonia | J10,J12-J18, J86 |

**Table S2: Top ten hits associated with the HMOX1 repeat.**

| **Phenotype** | **No of cases** | **Beta** | **95% CI** | **FDR adjusted p** |
| --- | --- | --- | --- | --- |
| Operative procedures - main OPCS4: X71.4 Procurement of drugs for chemotherapy for neoplasm for regimens in Band 9 | 1867 | -0.09 | (-0.14--0.05) | 0.335 |
| Operative procedures - OPCS4: X71.4 Procurement of drugs for chemotherapy for neoplasm for regimens in Band 9 | 1910 | -0.09 | (-0.13--0.04) | 0.335 |
| Job coding: property/housing/land manager | 411 | 0.19 | (0.09-0.28) | 0.335 |
| Date N40 first reported (hyperplasia of prostate) | 30226 | -0.02 | (-0.04--0.01) | 0.349 |
| Workplace very dusty | 39158 | 0.02 | (0.01-0.04) | 0.349 |
| Operative procedures - OPCS4: S02.1 Abdominoplasty | 265 | 0.22 | (0.1-0.33) | 0.349 |
| Job code - historical: Property, housing and land managers | 438 | 0.17 | (0.07-0.26) | 0.349 |
| Operative procedures - secondary OPCS4: T67.6 Primary simple repair of tendon | 529 | -0.16 | (-0.24--0.07) | 0.349 |
| Treatment/medication code: calceos chewable tablet | 247 | 0.22 | (0.1-0.34) | 0.349 |
| Treatment/medication code: otomize ear spray | 220 | 0.23 | (0.1-0.36) | 0.34 |

**Figure S1: Absolute difference in repeat length and true repeat length across all four cohorts.**

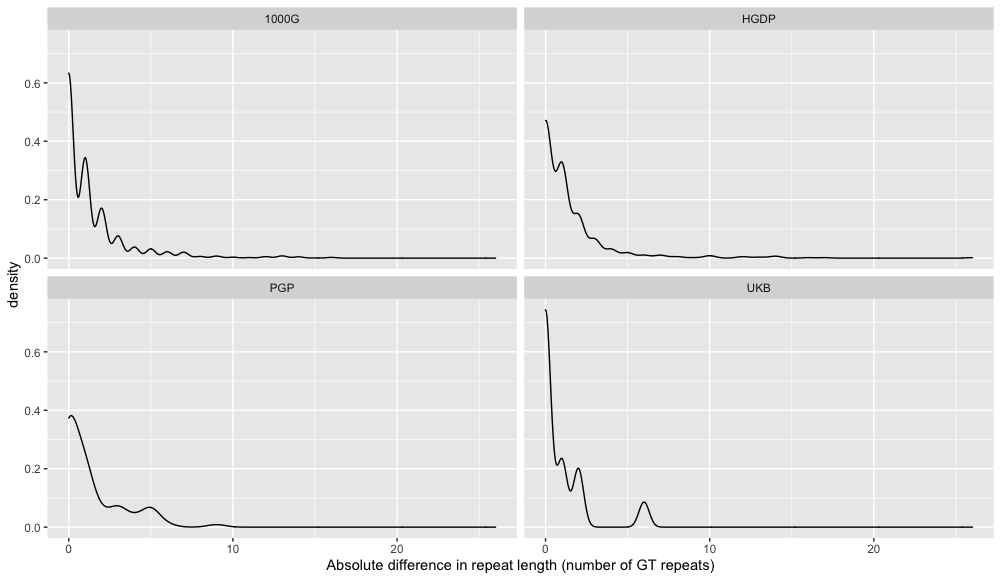
