## Supplement S2 for "Phenotypic associations with the *HMOX1* GT(n) promoter repeat in European populations"

Supplement S2: Imputation testing and quality control.

Background and WGS genotype calling:

The accuracy of the imputation of the HMOX1 STR from SNP data is critical to downstream analysis. Previously, Saini and colleagues had developed a haplotype reference panel for a wide range of STR’s and tested it on the 1000 Genomes project and Simon’s Simplex Collection (SSC) .^1^ As 1000 Genomes and SSC have ~ 30X WGS data, the STR can be called reliably, and the quality of the imputation compared with the SNP called imputation. Using this data, they reported reasonable quality imputation in European populations and in SSC but weaker performance in other populations.

The summary Pearson’s correlation in European populations was 0.97 and in SSC (largely European) the corresponding figure was 0.93. To confirm these figures, we replicated the analysis in four other cohorts with linked WGS data where the microsatellite length can be accurately called – The Human Genome Diversity Project (HGDP, n = 826),^2^ UK Biobank (UKB, n = 44), the UK Personal Genome Project (PGP, n = 88), and the 1000 Genomes project (1000G, n = 2318).

For HGDP, 1000G, and PGP, genetic data is freely available and the quality control on data is described in the relevant publication. For UKB, we utilised genetic data which had undergone in house quality control as described here.^3^ At the time of publication, only a pilot sample of UKB participants had undergone whole genome sequencing, hence the low numbers of included participants (n = 44).

For HGDP, UKB, and PGP, we downloaded BAM files from the respective cohorts, and used HipSTR v0.62 with standard settings to measure the STR repeat length. This uses a hidden Markov model to genotype and phase short tandem repeats while taking account of sequencing artefact We then performed the standard recommended filtering of genotype calls, filtering on the posterior probability (>0.9), and other technical aspects of the STR calling process as described here.

For 1000G, we contacted the authors of Saini et al, who kindly provided their WGS STR genotype calls directly.^1^

HMOX1 STR imputation

Subsequently, we then performed imputation using the haplotype reference panel on SNP array data from each cohort, using Beagle 4.2 with standard settings. Each cohort was imputed separately. Table S2 describes the number of included cases for comparison. We imputed across 1mb either side of the HMOX1 repeat, after initial experimentation and published data found no beneficial effect of larger imputation windows.^1^

Alleles were called based on the most probable allele as reported by Beagle, with no filtering due to posterior genotype probability (discussed in section below).

Table S3: Number of samples with both SNP and array data available.

| Study | Total | Imputed (SNP) | WGS accurately | Both available |
| --- | --- | --- | --- | --- |
| 1000G | 2318 | 2318 | 2133 | 2133 |
| HGDP | 826 | 800 | 676 | 656 |
| PGP | 88 | 87 | 83 | 83 |
| UKB | 44 | 44 | 44 | 44 |

Comparison of WGS vs imputed repeated length

For our initial analysis, we can simply compare the imputed repeat length (total) vs the WGS called repeat length (total). As described in detail in other sections, as HipSTR calling cannot phase a genotype (i.e. know which strand a read has come from), we have to assess the accuracy of the imputation at the genotype level, not the allelic level. All lengths were measured as the number of GT repeats.

We chose three metrics: Pearson’s correlation, which was simply calculated across the total allele length, exact concordance (total repeat length exactly matches the imputed length), and within two repeats (to identify the proportion of total lengths that were close to the exact length).

**Table S4: Metrics of imputation performance across each of the study cohorts**

| Study | Pearson’s correlation | Concordance (exact) | Within two repeats | n |
| --- | --- | --- | --- | --- |
| Overall | 0.907 | 0.455 | 0.837 | 2916 |
| 1000G | 0.914 | 0.460 | 0.835 | 2133 |
| HGDP | 0.876 | 0.426 | 0.848 | 656 |
| PGP | 0.900 | 0.469 | 0.771 | 83 |
| UKB | 0.936 | 0.590 | 0.932 | 44 |

As can be seen, across all four cohorts, the Pearson's correlation is > 0.85, and generally > 0.9, suggesting a very strong correlation between imputed and directly genotyped repeat length, backed up by the fraction of samples that have a total length within two repeats being > 0.80 in all but one cohort. However, the actual concordance - the number of repeat lengths that are exactly accurate, is lower. This is to be expected, as minor differences in repeat length in either allele will alter this. This can be seen visually in **Figure S2,** which plots all four studies on top of each other.

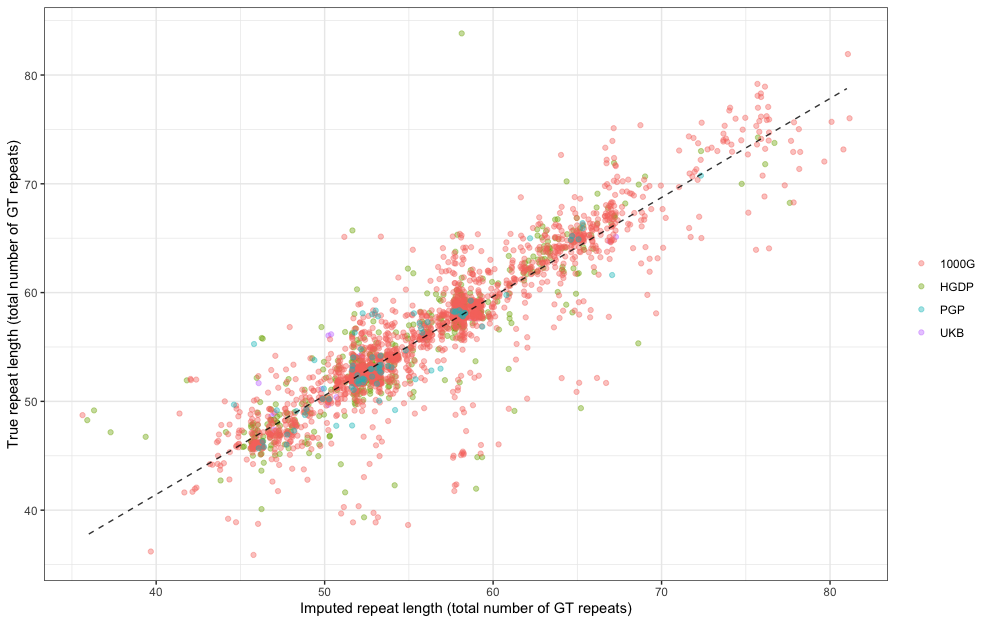

Geographic variation

It is likely that the imputation reference panel will perform differently in different populations. We therefore extracted superpopulation and population for the two cohorts where this was available (HGDP and 1000G).

In **Figure S3**, the imputation quality is plotted for each super population in HGDP against the number of participants. For all superpopulations except Oceania, the Pearson’s correlation was >0.7, but with evidence of varying imputation performance across each superpopulation.

Figure S3: Imputation quality across each superpopulation in HGDP.

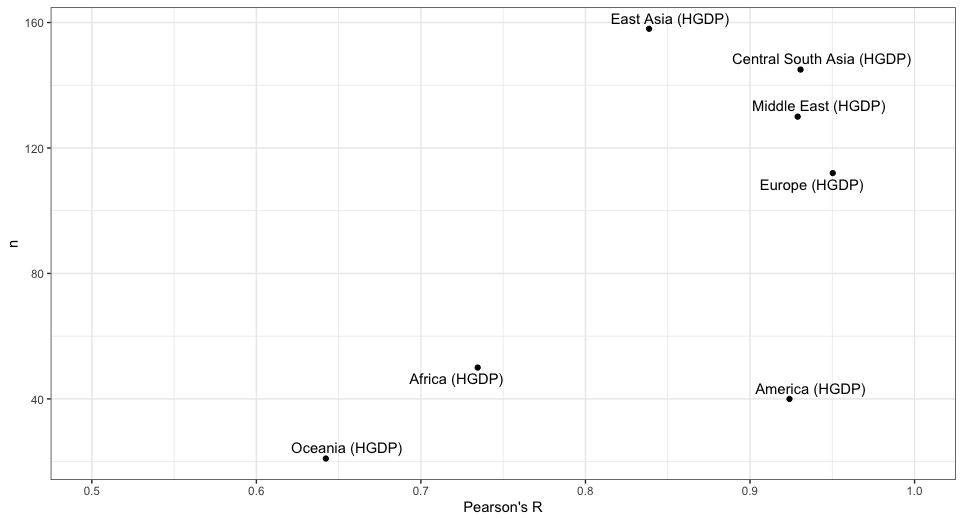

In **Figure S4,** we show the imputation performance across each population (n>10) in both 1000G and HGDP, again showing a large variation in imputation performance across sub populations. Table S3 describes the imputation performance for all three metrics within each population.

**Figure S4: Imputation performance across each population in HGDP and 1000G**

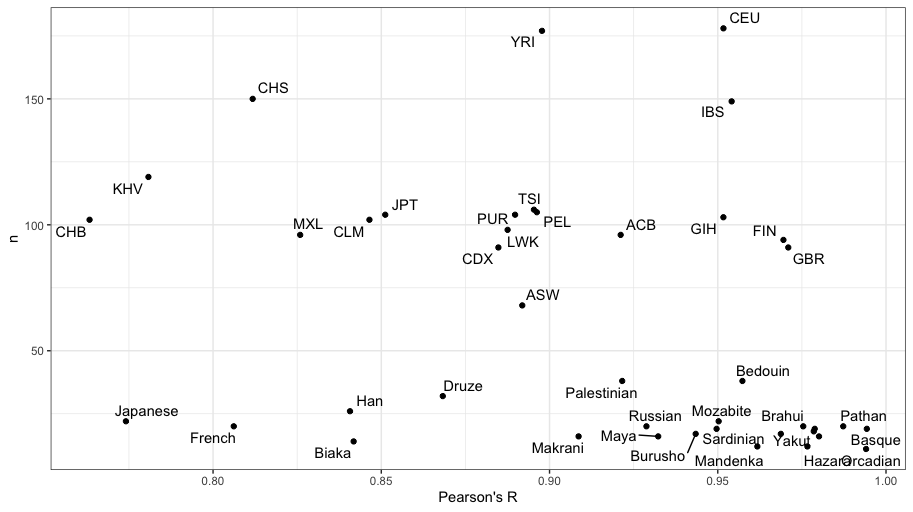

**Table S3: Imputation performance within each population in HGDP and 1000G**

| Population | Number of samples | Correlation | Concordance | Within 2 repeat lengths |
| --- | --- | --- | --- | --- |
| HGDP | | | | |
| Orcadian | 11 | 0.99 | 0.45 | 1.00 |
| Hazara | 12 | 0.98 | 0.50 | 0.92 |
| Mandenka | 12 | 0.96 | 0.17 | 0.67 |
| Biaka | 14 | 0.84 | 0.21 | 0.36 |
| Kalash | 16 | 0.98 | 0.50 | 0.88 |
| Makrani | 16 | 0.91 | 0.50 | 0.88 |
| Maya | 16 | 0.93 | 0.62 | 0.94 |
| Burusho | 17 | 0.94 | 0.47 | 0.88 |
| Yakut | 17 | 0.97 | 0.41 | 0.94 |
| Sindhi | 18 | 0.98 | 0.39 | 0.89 |
| Balochi | 19 | 0.98 | 0.53 | 0.95 |
| Basque | 19 | 0.99 | 0.58 | 1.00 |
| Sardinian | 19 | 0.95 | 0.37 | 0.95 |
| Brahui | 20 | 0.98 | 0.65 | 0.90 |
| French | 20 | 0.81 | 0.55 | 0.80 |
| Pathan | 20 | 0.99 | 0.65 | 1.00 |
| Russian | 20 | 0.93 | 0.65 | 0.90 |
| Japanese | 22 | 0.77 | 0.23 | 0.82 |
| Mozabite | 22 | 0.95 | 0.50 | 0.86 |
| Han | 26 | 0.84 | 0.23 | 0.77 |
| Druze | 32 | 0.87 | 0.38 | 0.84 |
| Bedouin | 38 | 0.96 | 0.53 | 0.87 |
| Palestinian | 38 | 0.92 | 0.42 | 0.87 |
| 1000G | | | | |
| ASW | 68 | 0.89 | 0.37 | 0.72 |
| CDX | 91 | 0.88 | 0.36 | 0.81 |
| GBR | 91 | 0.97 | 0.67 | 0.93 |
| FIN | 94 | 0.97 | 0.74 | 0.91 |
| ACB | 96 | 0.92 | 0.32 | 0.68 |
| MXL | 96 | 0.83 | 0.55 | 0.91 |
| LWK | 98 | 0.89 | 0.19 | 0.66 |
| CHB | 102 | 0.76 | 0.38 | 0.86 |
| CLM | 102 | 0.85 | 0.54 | 0.93 |
| GIH | 103 | 0.95 | 0.31 | 0.87 |
| JPT | 104 | 0.85 | 0.30 | 0.76 |
| PUR | 104 | 0.89 | 0.46 | 0.82 |
| PEL | 105 | 0.90 | 0.69 | 0.96 |
| TSI | 106 | 0.90 | 0.57 | 0.86 |
| KHV | 119 | 0.78 | 0.29 | 0.78 |
| NA | 127 | 0.91 | 0.51 | 0.83 |
| IBS | 149 | 0.95 | 0.63 | 0.90 |
| CHS | 150 | 0.81 | 0.41 | 0.85 |
| YRI | 177 | 0.90 | 0.35 | 0.73 |
| CEU | 178 | 0.95 | 0.58 | 0.89 |

In summary, imputation performance was reasonable for identifying whether the total repeat length across both alleles was similar. Exact concordance was less likely, but the proportion of samples where the total repeat length was more than 2 repeats away was <20%.

Impact of filtering post genotype

As Beagle computes posterior probabilities for each called genotype, we perform some further testing to measure whether the computed posterior probability was useful in filtering low quality calls. As Beagle is generally used to call biallelic variants, it was not known whether the reported genotype posterior probability would have relevance in a multiallelic (64 separate STR length) setting.

To do this, we used the 1000G data to test whether the reported posterior probability at each genotype related to the probability of that genotype being correct. **Figure S5A** shows the histogram posterior probability for each reported genotype. The vast majority (79.3%) of reported posterior probabilities were one, with the next commonest reported probabilities being 0.99 (3.6%) and 0.98 (1.86%). Despite this, there was almost no correlation between the reported posterior probability and absolute mean difference in repeat length (**Figure S5B**). There was a slight correlation between the reported posterior probability and repeat length, with longer repeats more likely to have higher posterior probabilities (**Figure S5C**). For completeness, we then re-ran the analysis calculating repeat length based on the average repeat length summed across all posterior probabilities. i.e., for a sample that had an estimated repeat length of 48 (posterior probability 0.8) and 42 (posterior probability 0.2), we would calculate the repeat length as 48 * 0.8 + 42 * 0.2 = 46.8 This approach is shown in **Figure S5D,** making clear that the addition of this methodology makes little difference (as would be expected, with most genotypes being called with >95% reported probability).

Figure S5: Effect of filtration on accuracy of genotype calls: A) Histogram of reported posterior probabilities, B) Posterior probability plotted against absolute difference in repeats, C) posterior probability plotted against repeat length, and D) Comparison of taking the most probable allele vs the full posterior approach.

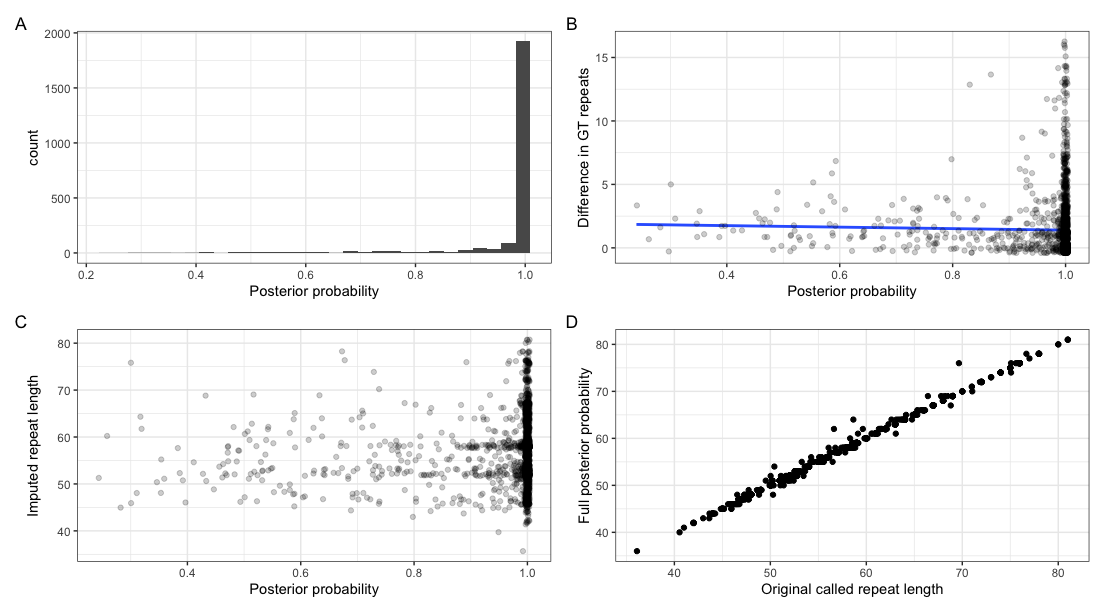

1.	Saini S, Mitra I, Mousavi N, Fotsing SF, Gymrek M. A reference haplotype panel for genome-wide imputation of short tandem repeats. Nat Commun [Internet] 2018;9(1):4397. Available from: http://dx.doi.org/10.1038/s41467-018-06694-0

2.	Cavalli-Sforza LL. The Human Genome Diversity Project: past, present and future. Nat Rev Genet [Internet] 2005;6(4):333–40. Available from: http://dx.doi.org/10.1038/nrg1596

3.	Mitchell R, Hemani G, Dudding T, Corbin L, Harrison S, Paternoster L. UK Biobank Genetic Data: MRC-IEU Quality Control, version 2. 2019 [cited 2021 Dec 21];Available from: https://data.bris.ac.uk/data/dataset/1ovaau5sxunp2cv8rcy88688v
